# Neutrophil-to-Lymphocyte Ratio Predicts Infusion-Site Skin Nodules in Parkinson’s Disease Patients Receiving Foslevodopa/Foscarbidopa Subcutaneous Infusion

**DOI:** 10.64898/2026.03.24.26349214

**Authors:** Elena Contaldi, Luca Magistrelli, Silvia Piazza, Arturo Caniglia, Ennio A. Mainardi, Pasquale Giametta, Gianni Pezzoli, Ioannis U. Isaias, Giulia Lazzeri

## Abstract

**Background:** Continuous subcutaneous foslevodopa/foscarbidopa infusion (LDp/CDp-CSI) is an effective treatment for patients with Parkinson’s disease (PD), but infusion-site nodules are a major cause of treatment discontinuation. Systemic inflammation can influence local skin tolerance; however, predictive biomarkers remain unidentified.

**Objective:** To evaluate the predictive value of the neutrophil-to-lymphocyte ratio (NLR) for clinically significant infusion-site nodules (PD-CSN) during LDp/CDp-CSI and to establish a clinical management framework to mitigate their development.

**Methods:** We prospectively followed 38 patients with PD initiating LDp/CDp-CSI for ≥3 months. Baseline immunological data were collected before infusion. A subset of 30 patients was followed for an average of 11 months to identify factors associated with skin nodules at longer follow-up. Nodules were classified by blinded raters. Between-group comparisons, ANCOVA, ROC curve, and Kaplan-Meier analyses were performed.

**Results:** At 3 months, 42% of patients were PD-CSN and showed higher baseline neutrophil counts (*P*=0.030) and NLR (*P*=0.007), with NLR remaining independently associated with nodule status (F=7.06, *P*=0.012). ROC analysis demonstrated acceptable discrimination (AUC=0.73, *P*=0.016). At last follow-up, lower baseline lymphocyte counts (*P*=0.002) and higher NLR (*P*=0.001) were observed in PD-CSN. High baseline NLR predicted earlier nodule onset (*P*=0.001). Despite frequent nodules, multidisciplinary team surveillance, including remote and in-person follow-up, limited treatment discontinuation to 5.3%.

**Conclusions:** Baseline systemic inflammation, reflected by NLR, predicts both the onset and persistence of infusion-site nodules during LDp/CDp-CSI. NLR may serve as a clinically accessible biomarker for early risk stratification. Multidisciplinary surveillance facilitates timely nodule management and enhances treatment adherence.

## Introduction

Levodopa remains the main symptomatic therapy for patients with Parkinson’s disease (PD), yet its use can lead to motor complications, including motor fluctuations and levodopa-induced dyskinesias [1]. Motor fluctuations affect up to 50% of PD patients within five years, significantly reducing quality of life [2]. For advanced PD patients refractory to optimized oral therapy or ineligible for other device-aided treatments, continuous subcutaneous foslevodopa/foscarbidopa infusion (LDp/CDp-CSI) provides steady dopaminergic delivery and improves “on” time compared to oral therapy. However, infusion-site reactions such as erythema, cellulitis, oedema, and nodule formation are common and may lead to discontinuation in up to 25% of patients [3, 4]. Algorithms for the management and prevention of cutaneous side effects have been developed to improve quality of life and limit the interruption of subcutaneous therapy [5]. In particular, meticulous hygiene of the needle insertion site, regular skin care, rotation of insertion sites, daily or alternate-day needle changes, and topical corticosteroid treatment have been identified as effective management strategies [6].

Histologically, nodules show local neutrophilic and lymphocytic infiltration, reflecting chronic inflammation [7, 8]. In this context, it is conceivable that systemic inflammatory activity contributes to altered local skin tolerance; however, the predictive role of inflammatory biomarkers remains largely unexplored. The neutrophil-to-lymphocyte ratio (NLR) is a readily available marker of systemic inflammation, previously associated with PD [9–11]. We hypothesized that elevated baseline NLR may predispose patients to clinically significant infusion-site nodules. This study aimed to assess the association between baseline NLR and nodule occurrence in real-world PD patients treated with LDp/CDp-CSI and to identify factors influencing infusion tolerability and treatment discontinuation. In parallel, we introduced an integrated care model designed to support long-term treatment adherence. This structured, multidisciplinary approach, incorporating standardized skin-care procedures, proactive patient education, and regular in-person and remote monitoring, facilitated early recognition and timely management of infusion-site events during LDp/CDp-CSI therapy.

## Methods

### Study Population and Baseline Assessment

We conducted a prospective observational study at the Parkinson Institute of Milan (ASST Gaetano Pini-CTO). Patients initiating LDp/CDp-CSI were consecutively enrolled between February 2024 and June 2025. Inclusion criteria were the following: i) a diagnosis of idiopathic PD according to current criteria [12]; ii) initiation of LDp/CDp-CSI with at least one documented follow-up assessment; and iii) availability of complete baseline data for inflammatory markers. As exclusion criteria, we considered: i) any concurrent acute inflammatory or infectious disease at enrollment; ii) a history of chronic autoimmune diseases or malignancy; iii) current or recent (<3 months) use of systemic immunosuppressive or immunomodulatory therapies (e.g., corticosteroids, biologics, or cytotoxic agents) that could markedly alter inflammatory status. Study procedures were approved by the local Ethics Committee (protocol 3499/23), and this research was carried out in agreement with the principles of the Declaration of Helsinki and later amendments. All patients gave their written informed consent before inclusion in the study.

Before initiating LDp/CDp-CSI, patients underwent a comprehensive clinical evaluation and confirmation of all inclusion and exclusion criteria. Baseline assessments included motor evaluation in medication “on” condition (Unified Parkinson’s Disease Rating Scale [UPDRS] part III and Hoehn & Yahr (HY) scale [13, 14]) and a brief cognitive screening (Mini-Mental State Examination [MMSE [15]], Montreal Cognitive Assessment [MoCA [16]], and Frontal Assessment Battery [FAB [17]], with scores adjusted for age and education level). Peripheral venous blood sampling for baseline NLR determination (neutrophil count divided by lymphocyte count) was performed, followed by the transition from prior dopaminergic therapy and the initiation of LDp/CDp-CSI under medical supervision. LDp/CDp-CSI dose titration and optimization of infusion parameters required three to five days of inpatient care and were combined with structured patient and caregiver education on infusion management. Training covered device handling and troubleshooting, infusion-site care (including systematic rotation of subcutaneous infusion sites across abdominal quadrants and strict adherence to aseptic technique), and routine cannula replacement every 24 hours to optimize local site management. Cannula length (6 mm or 9 mm) was selected according to abdominal subcutaneous tissue thickness, with 9-mm cannulas preferentially used in patients with more pronounced adipose tissue. The replacement interval was subsequently individualized based on patient tolerability during follow-up assessments. Patients were also instructed to apply a gentle local massage around the infusion area.

### Follow-up Schedules and Nodule Evaluation

Following in-hospital LDp/CDp-CSI titration, patients were re-evaluated within the framework of a dedicated outpatient care protocol (Italian Modello Ambulatoriale Complesso [MAC]) involving both neurologists and nurse specialists. Scheduled assessments were conducted at fixed time points (three, six, and twelve months ±1 month) and additionally as needed according to individual clinical requirements. At each evaluation, subcutaneous nodules were systematically and independently assessed by a nurse specialist (S.P.) and a neurologist (G.L.), who were blinded to baseline inflammatory markers.

According to a classification adapted from Todd and James [18], nodules were categorized based on: i) size and induration, ii) presence and intensity of erythema or other inflammatory signs, iii) associated symptoms such as pain, and iv) interference with daily activities or need for medical intervention. Mild nodules were defined as small (<1 cm), non-tender, or minimally inflamed lesions that did not interfere with infusion.

Moderate nodules showed greater enlargement (1-3 cm) associated with clear inflammatory changes, occasionally accompanied by local discomfort. Severe nodules were larger (>3 cm), painful, inflamed, or functionally limiting, often necessitating medical treatment. For the present study, this three-level classification was simplified into a binary outcome (clinically significant nodules, PD-CSN; no significant reactions, PD-NSR). Patients were instructed to promptly report new or worsening nodules via email or a dedicated phone line, and a nurse specialist provided guidance and rapidly assessed events, ensuring close monitoring between in-person visits. Mild skin reactions were managed with topical formulations stimulating fibroblast activity and promoting tissue regeneration. Moderate nodules were treated with topical corticosteroids and monitored through a structured remote protocol, including photographic documentation sent via email or phone, with additional in-person evaluation scheduled within a few days whenever needed. Alternative infusion sites (including the arm) were considered when abdominal locations were poorly tolerated. Severe nodules with marked inflammatory signs were managed with oral antibiotic therapy under medical supervision. Time-to-event data were prospectively collected for all clinically significant nodules.

The nurse specialist also assessed patient or caregiver adherence to infusion-site management procedures, and these data were incorporated into the comparative analysis between PD-CSN and PD-NSR.

### Statistical Analyses

This is a preliminary hypothesis-generating longitudinal study; therefore, formal sample size estimation was not calculated.

Variables were expressed as counts (percentages) when categorical and as means (standard deviation, SD) when continuous. The normality of data was assessed using the Shapiro-Wilk test. Inter-rater reliability between the nurse specialist and the neurologist for nodule classification was evaluated through Cohen’s kappa as follows: values ≤ 0 indicating no agreement, 0.01–0.20 as none to slight, 0.21–0.40 as fair, 0.41–0.60 as moderate, 0.61–0.80 as substantial, and 0.81–1.00 as almost perfect agreement [19]. If there was disagreement, a third rater (L.M.) was involved. The final classification used for analysis was based on the consensus, including the third evaluator’s assessment.

Differences between groups were analyzed by an independent samples t-test or the non-parametric equivalent Mann-Whitney test, as appropriate. Categorical variables were analyzed using the Chi-square test or, when appropriate, Fisher’s exact test. Post hoc power analysis was performed based on the primary outcome (difference in NLR between PD-CSN and PD-NSR at three months). Analysis of covariance (ANCOVA) was used to adjust for relevant covariates, as the limited number of PD-CSN cases precluded reliable multivariable regression modeling. Key demographic covariates were age and sex, known to strongly influence immunological measures [20], along with additional covariates selected based on between-group comparisons and Spearman correlation analyses. Exploratory models additionally included foslevodopa LEDD to examine potential pharmacological confounding. Model assumptions (linearity, homogeneity of regression slopes, normality of residuals, and absence of influential outliers) were verified and met. F-values, partial eta-squared (η□²), and corresponding *P*-values, were reported. A receiver operating characteristic (ROC) curve analysis was performed to explore biomarkers’ discriminatory power, obtaining the area under the curve (AUC) and significance values. AUC interpretation was determined according to Mandrekar et al. [21]. Finally, time-to-event analysis was performed to assess the association between NLR and the occurrence of CSN at the last follow-up. The probability of event-free time was estimated using the Kaplan-Meier (KM) method. Differences in event-free time curves between the two NLR groups (NLR-high *vs* NLR-low, dichotomized by the optimal cut-off) were compared using the log-rank test.

Two patients were receiving antiplatelet therapy (clopidogrel and aspirin), but immunological measures were within normal range limits. To assess the robustness of the results and account for the potential confounding effects of these medications, sensitivity analyses were also performed. All tests were two-tailed, and the significance level was *P* < 0.05. Analyses were performed using SPSS version 25 (IBM Corporation, Armonk, USA), RStudio 2022.07.2+576, GraphPad Prism version 8 (GraphPad Software Inc., San Diego, USA), and Jamovi (Version 2.3.28).

## Results

### Patient Selection and Baseline Characteristics

Of the 44 patients initially screened, six were excluded from the study. Three were excluded during initial screening for failing to meet the eligibility criteria: one had an intercurrent urinary tract infection with altered blood cell counts at the time of blood sampling, and two had concurrent autoimmune and hematologic diseases. An additional three patients were excluded during follow-up: two developed symptoms suggestive of atypical parkinsonian syndromes, and one was lost to follow-up. The final analyzed cohort comprised 38 patients (see Table 1 for complete baseline measures).

### Development of Clinically Significant Nodules at 3-month follow-up

Within 3 months, 16 patients (42%) developed clinically significant nodules. Inter-rater reliability for nodule classification was substantial (Cohen’s Kappa coefficient = 0.834, SE = 0.091, *P* <0.001). Disagreement occurred only in three observations, which were reviewed and resolved by the third rater.

Demographic and clinical characteristics were comparable between PD-CSN and PD-NSR, except for lower MoCA scores in PD-CSN. There were no significant differences in infusion-related parameters, see Table 2. Concerning immunological analyses, PD-CSN had higher neutrophil counts (3.44 ± 0.90 *vs* 2.85 ± 0.72×10□/L, *P* = 0.030) and higher NLR values (2.16 ± 0.61 *vs* 1.65 ± 0.48, *P* = 0.007), see Figure 1A-B. Post hoc power analysis indicated a Cohen’s d effect size of 0.94 and an achieved power of 0.81 (two-tailed α = 0.05).

**Figure 1:**
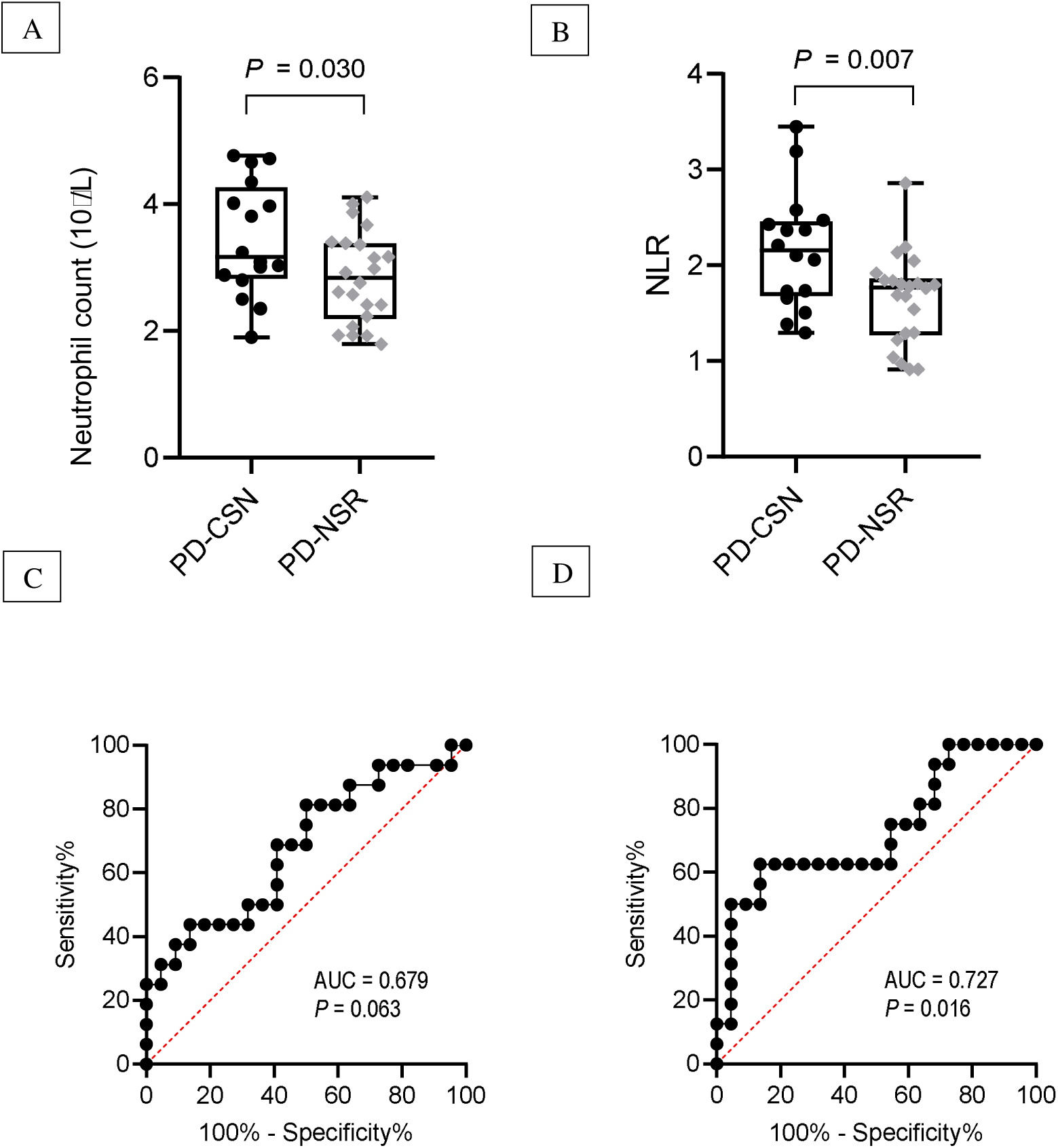
Comparison of mean neutrophil count (A) and neutrophil-to-lymphocyte ratio (NLR; B) between PD-CSN and PD-NSR groups at the 3-month follow-up. Receiver operating characteristic (ROC) curve analyses for neutrophil count (C) and NLR (D) in discriminating PD-CSN.

The principal ANCOVA model on N = 38, adjusted for age, sex, MoCA score, and BMI, showed that the fixed factor nodule status had a significant effect on neutrophil count (F = 10.856, η□² = 0.259, *P* = 0.002). A significant association was also observed for BMI (F = 9.479, η□² = 0.234, *P* = 0.004). Similar findings were confirmed in the sensitivity analysis on N = 36 (nodule status: F = 10.170, η□² = 0.260, *P* = 0.003; BMI: F = 8.441, η□² = 0.225, *P* = 0.007). An exploratory ANCOVA including nodule status, age, sex, BMI, MoCA, and foslevodopa LEDD at three months was fitted on the whole cohort (N = 38) to assess potential pharmacological confounding. In this full model, the association between neutrophil count and nodule status remained statistically significant (F = 9.092, η□² = 0.233, *P* = 0.005), supporting its independence from foslevodopa dose. When the same analyses were performed for NLR, the principal ANCOVA model (adjusted for age, sex, MoCA score, and BMI) showed that nodule status had a significant effect on the outcome (F = 7.063, η□² = 0.186, *P* = 0.012), which was confirmed in the sensitivity analysis (F = 4.241, η□² = 0.128, *P* = 0.049) on N = 36. In the exploratory full model, the association between nodule status and NLR remained statistically significant (F = 5.366, η□² = 0.152, *P* = 0.028). Results are summarized in Table 3.

ROC analysis confirmed acceptable discrimination of PD-CSN for NLR (AUC = 0.727, 95% CI: 0.557-0.896, *P =* 0.016) but not for neutrophil count (AUC = 0.679, *P* = 0.063), Figure 1C-D. Similar results were found in the sensitivity analysis (Supplementary Table S1). An optimal cut-off ≥2.05 showed 62.5% sensitivity and 86.4% specificity in detecting PD-CSN. Based on this value, two distinct groups (NLR high, NLR low) were identified.

### Presence of Clinically Significant Nodules at Last Available Follow-up

Among 30 patients with a follow-up of ≥6 months, 12 (40%) had clinically significant nodules, with a mean time to onset of 3.5 months. Ten patients (83%) had persistent nodules at both assessments, while two developed new nodules and three showed resolution of previously present nodules. Notably, only two patients (5.3% of the total cohort) discontinued LDp/CDp-CSI due to a subjective lack of efficacy (one PD-CSN and one PD-NSR). Adherence to the standardized management protocol was high, reflecting the effectiveness of structured patient and caregiver training during inpatient care and subsequent outpatient follow-up. Patients and caregivers demonstrated competence in device handling and infusion management (81.25% of PD-CSN *vs* 86.36% of PD-NSR, *P* = 0.682).

PD-CSN had significantly lower BMI and FAB than PD-NSR, see Table 4. Furthermore, PD-CSN showed lower lymphocyte count (1.38 ± 0.31 *vs* 1.97 ± 0.50×10□/L, *P* = 0.002) and higher NLR (2.35 ± 0.53 *vs* 1.60 *±* 0.51, *P* = 0.001), Figure 2A-B.

**Figure 2:**
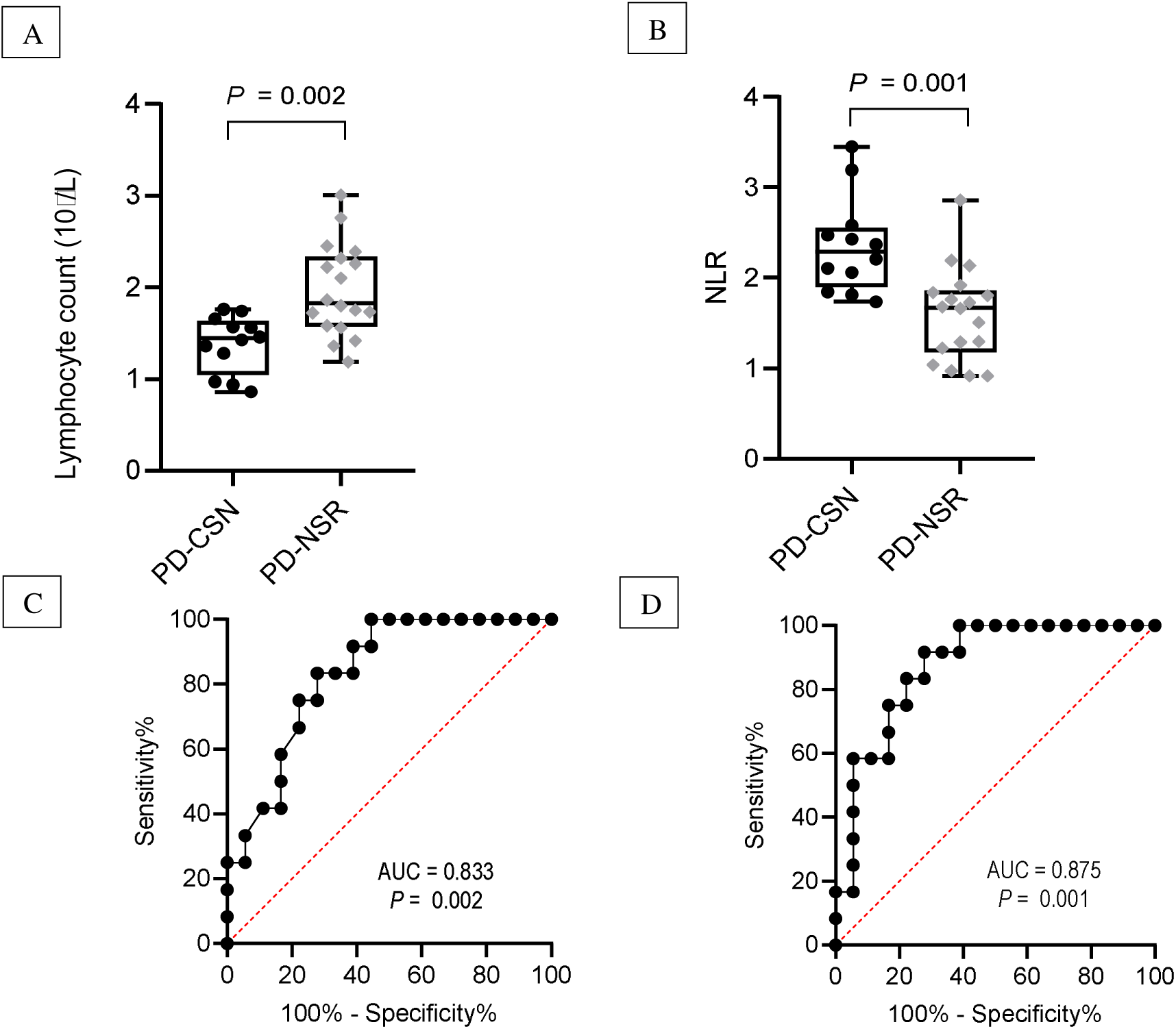
Comparison of mean lymphocyte count (A) and neutrophil-to-lymphocyte ratio (NLR; B) between PD-CSN and PD-NSR groups at the last follow-up. Receiver operating characteristic (ROC) curve analyses for lymphocyte count (C) and NLR (D) in discriminating PD-CSN.

Due to sample size constraints, distinct principal ANCOVA models were employed on N = 30.

Model 1, adjusted for age, sex, and BMI, showed that nodule status had a significant effect on lymphocyte count (F = 6.773, η□² = 0.220, *P* = 0.016), with trends confirmed in the sensitivity analysis on N = 28 (*P* = 0.073). Model 2, adjusted for age, sex, and FAB score, also showed a significant effect of nodule status on lymphocyte count (F = 8.753, η□² = 0.267, *P* = 0.007), with significance confirmed in the sensitivity analysis (F = 6.424, η□² = 0.234, *P* = 0.019). Distinct exploratory ANCOVA models were then performed to assess pharmacological confounding. In Model 1 (adjusted for age, sex, BMI, and foslevodopa LEDD at last follow-up), there was a significant effect of nodule status (F = 5.538, η□² = 0.187, *P* = 0.027). In Model 2 (adjusted for age, sex, FAB score, and foslevodopa LEDD), the effect of nodule status remained significant (F = 8.378, η□² = 0.267, *P* = 0.008). When the same analyses were repeated for NLR, Model 1 (adjusted for age, sex, and BMI) showed a significant effect of nodule status (F = 18.168, η□² = 0.431, *P* < 0.001), which was confirmed in the sensitivity analysis (F = 12.725, η□² = 0.356, *P* = 0.002). Model 2 (adjusted for age, sex, and FAB score) also showed a significant effect (F = 12.694, η□² = 0.346, *P* = 0.002), confirmed in the sensitivity analysis (F = 8.989, η□² = 0.290, *P* = 0.007). Exploratory models including foslevodopa LEDD confirmed consistent results (Model 1: F = 15.518, η□² = 0.403, *P* = 0.001; Model 2: F = 13.837, η□² = 0.376, *P* = 0.001). Results are summarized in Table 3.

Both lymphocyte count and NLR were able to discriminate PD-CSN in ROC curve analysis (AUC = 0.833, 95% 0.691-0.975, *P* = 0.002; AUC = 0.875, 95% CI = 0.751-0.995, *P* = 0.001; Figure 2C-D); however, with better performance of NLR. AUC and *P*-values were similar in the sensitivity analysis (Supplementary Table S1).

### Survival analysis

An exploratory KM model was employed to detect differences between the NLR high and NLR low groups. In the NLR low group (N = 24), 20.83% experienced the event *vs* 71.43% in the NLR high group (N = 14). The time to nodule development was significantly longer for the NLR low group at 14.22 months compared to the NLR high group at 6 months. The difference was significant (log-rank χ² = 10.411, *P* = 0.001; Supplementary Figure 1) and consistent in sensitivity analyses (χ^2^ = 8.481, *P* = 0.004).

## Discussion

In this study, we explored clinical and immunological factors associated with clinically relevant nodules in PD patients receiving LDp/CDp-CSI. Our findings can be summarized as follows: i) clinically significant nodules developed in 42% of patients within the first three months, with a largely stable pattern over time; ii) PD-CSN had higher baseline neutrophil counts and NLR than PD-NSR, independent of relevant clinical and demographic factors; iii) at later follow-up, PD-CSN also exhibited lower lymphocyte counts alongside higher NLR; iv) survival analyses confirmed that higher baseline NLR predicted earlier nodule onset.

Collectively, these data suggest that systemic inflammatory status may predispose to local cutaneous intolerance during LDp/CDp-CSI. It is also interesting to highlight that, though nodules were relatively common, the discontinuation rate of infusion therapy in our series was very low (5.3L%), markedly below the rates reported in registrational and earlier realLworld cohorts.

When contextualizing our data within the available clinical evidence, phase 3 randomized controlled trials of LDp/CDp-CSI reported discontinuation rates due to adverse events of 22% in the treatment arm compared with 1% in the oral levodopa-carbidopa group [3]. In a 52-week open-label extension involving 244 patients, infusion-site complications were the most common adverse events, leading to treatment discontinuation in nearly 44% of participants [22]. Real-world data have been equally heterogeneous. Rukavina et al. observed skin reactions in 31% of patients and a discontinuation rate of 24.5% [4], whereas another study of 24 patients reported treatment cessation in 40%, predominantly attributable to inflammatory skin reactions at abdominal infusion sites, which were observed in half of the cohort [23]. Similar rates of therapy interruption were detected in a series of 26 patients (30.77%) [24], with six patients reporting insufficient efficacy and only one developing cellulitis. Only a phase I study documented a substantially lower discontinuation rate, in line with our observations (1/15 patients, approximately 6.7%), although the evaluation period in that study was limited to 72 hours of LDp/CDp-CSI [25]. In this context, the low discontinuation rate observed in our cohort represents a relevant distinguishing feature, likely reflecting the implementation of a standardized management protocol provided by neurologists and nurse specialists through both in-person and remote assessments. Notably, although clinically significant skin reactions occurred in approximately 40% of participants, the management by the dedicated multidisciplinary care team enabled early detection and timely treatment of infusion-site nodules, contributing to the very low overall discontinuation rate.

The future incorporation of additional biomarkers, such as NLR, may enable improved stratification of patients at risk for adverse events such as cutaneous lesions, allowing for optimized resource allocation and even closer monitoring of selected individuals. NLR represents a promptly available indicator of peripheral inflammation derived from two distinct but complementary leukocyte subpopulations [26], and has been largely investigated in PD [26–28]. Our findings expand this framework, suggesting that different immune-cell compartments may underlie distinct phases of the inflammatory response, from initial neutrophil activation to sustained lymphocyte-mediated dysregulation.

Neutrophils perform several functions, including the production of reactive oxygen species, phagocytosis, degranulation, and the release of neutrophilic extracellular traps. They are also involved in local tissue repair processes for their ability to eliminate pathogens [29], but an excessive activation may result in a prolonged inflammatory state [30]. Over time, particularly in older or immunosenescent individuals, impaired lymphocyte-mediated regulation may also impair the resolution of inflammation, leading to persistent nodules. Reduced lymphocyte counts have already been reported in PD [31] and reflect immunosenescence and “inflammaging”, a condition characterized by chronic low-grade inflammation and reduced immune competence [32]. The consequences of these mechanisms are well known at the central nervous system level, where impaired lymphocyte subsets may trigger neurotoxicity and neurodegeneration [33]. Peripherally, tissue-selective localization of lymphocytes to the skin is crucial for immune surveillance by promoting the maintenance of homeostasis [29]. In patients with marked lymphocytic dysregulation and continuous subcutaneous exposure, these mechanisms may be compromised, favoring the persistence and chronicization of skin reactions [34]. Interestingly, this distinct pattern of immune involvement mirrors histopathological observations of LDp/CDp infusion-site nodules, showing both neutrophilic and lymphocytic infiltrations [7, 8].

Among the clinical determinants of skin reactions, cognitive fragility (reflected by lower MoCA and FAB scores) and lower BMI were more frequent in PD-CSN. Cognitive deficits may predispose to nodule occurrence by compromising adherence to specific hygiene protocols, even under caregiver supervision. Alternatively, cognitive impairment in PD has been associated with immune dysregulation [26], suggesting that cognitive and cutaneous vulnerabilities could represent parallel manifestations of a more reactive and dysfunctional immune phenotype. Moreover, lower BMI may promote deeper tissue penetration and greater mechanical stress at infusion sites, a phenomenon also reported with continuous insulin infusion [35].

Importantly, even after adjusting for these potential confounding factors in ANCOVA analyses, the association between elevated NLR and skin nodule occurrence remained statistically significant, highlighting its potential role as an independent biological correlate of infusion-site pathology.

Strengths of this study include: i) prospective design, which allowed a careful assessment of both short- and medium-term factors associated with clinically relevant skin reactions; ii) systematic and blinded nodule assessment with high inter-rater reliability; iii) use of a standardized protocol of skin management and high-quality surveillance, which ensured robust and consistent event reporting; iv) integration of different statistical approaches. Several limitations should also be highlighted: i) relatively small sample size, reflecting the recent implementation of LDp/CDp-CSI in European clinical practice; ii) lack of histopathological assessment; iii) possible residual confounding despite thorough sensitivity analyses; iv) while the binary classification of nodules increases clinical interpretability, future studies could benefit from quantitative measures of lesion size and severity.

### Conclusions

Our findings indicate that systemic inflammatory dysregulation, reflected by elevated baseline NLR, increases susceptibility to both the onset and persistence of infusion-site skin reactions during LDp/CDp-CSI. Despite the underlying immunological involvement, coordinated multidisciplinary care ensured a low discontinuation rate. Given its simplicity and clinical accessibility, NLR may serve as a practical biomarker for early risk stratification and targeted preventive strategies. Future studies integrating peripheral cytokine profiling, immune cell phenotyping, and skin histology are needed to validate these observations and support more individualized management approaches.

## Data Availability

All data produced in the present study are available upon reasonable request to the authors

## Acknowledgments

The authors thank all patients and families for their contribution, as well as the Associazione Italiana Parkinsoniani and the Fondazione Pezzoli per la Malattia di Parkinson (Milan, Italy) for the long-lasting support.

## Author contributions

Conceptualization: E.C., G.L.; Statistical analyses: E.C.; Investigation: E.C., L.M., G.L.; Data Curation: S.P., A.C.; Writing of the original draft: E.C.; Review and editing: L.M., E.A.M., P.G., G.P., I.U.I., G.L.; Supervision: I.U.I.

## Statements and declarations

### Ethical considerations

Study procedures were approved by the local Ethics Committee (Comitato Etico Territoriale Lombardia 3, protocol 3499/23), and this research was carried out in agreement with the principles of the Declaration of Helsinki and later amendments.

### Consent to Participate

All participants provided written informed consent before inclusion in the study.

### Consent for publication

Not applicable.

### Conflict of Interest

IUI received research grants, speaking and consultant honoraria from Medtronic, Boston Scientific, Newronika, and the “Fondazione Pezzoli per la Malattia di Parkinson”, and speaking honoraria from AbbVie. The other authors have no conflict of interest to report.

### Funding statement

This work was supported by the unconditional and independent contribution of the ‘Fondazione Pezzoli per la Malattia di Parkinson’, Milan (Italy), (Italian ‘5x1000’ funding), a Research Foundation linked to the ‘Associazione Italiana Parkinsoniani’ (AIP).

### Data Availability Statement

The source data used for analyses presented in this study are available from the authors upon reasonable request.

