## Supplementary Material for "Neutrophil-to-Lymphocyte Ratio Predicts Infusion-Site Skin Nodules in Parkinson’s Disease Patients Receiving Foslevodopa/Foscarbidopa Subcutaneous Infusion"

c) Medical Management Unit, ASST G. Pini-CTO, Milan, Italy

d) Pezzoli Foundation for Parkinson’s Disease, Milan, Italy

e) Department of Neurology, University Hospital of Würzburg and Julius-Maximilian-University of Würzburg, Würzburg, Germany

**Supplementary Tables**

**Supplementary Table S1:** Sensitivity analysis of ROC curves for the detection of PD-CSN at 3 months and the last available follow-up by immunological measures.

ROC curve for PD-CSN at 3 months (N = 36)

Neutrophil count

AUC = 0.648 (95% CI: 0.461-0.834), *P* = 0.136

Neutrophil-to-lymphocyte ratio

AUC = 0.698 (95% CI: 0.515-0.882), *P* = 0.045

ROC curve for PD-CSN at last follow-up (N = 28)

Lymphocyte count

AUC = 0.826 (95% CI: 0.676-0.976), *P* = 0.004

Neutrophil-to-lymphocyte ratio

AUC = 0.861 (95% CI: 0.726-0.995), *P* = 0.001

**Supplementary Figures**

**Supplementary Figure 1:** Kaplan–Meier curves showing time to nodule development according to baseline NLR levels. Patients were divided into two groups based on the ROC curve cut-off (NLR high, ≥2.05; NLR low, <2.05). The y-axis represents the probability of remaining nodule-free over time, and censored observations are indicated by tick marks. The NLR high group showed a higher incidence and earlier onset of nodules compared with the NLR low group (log-rank test *P* = 0.001).

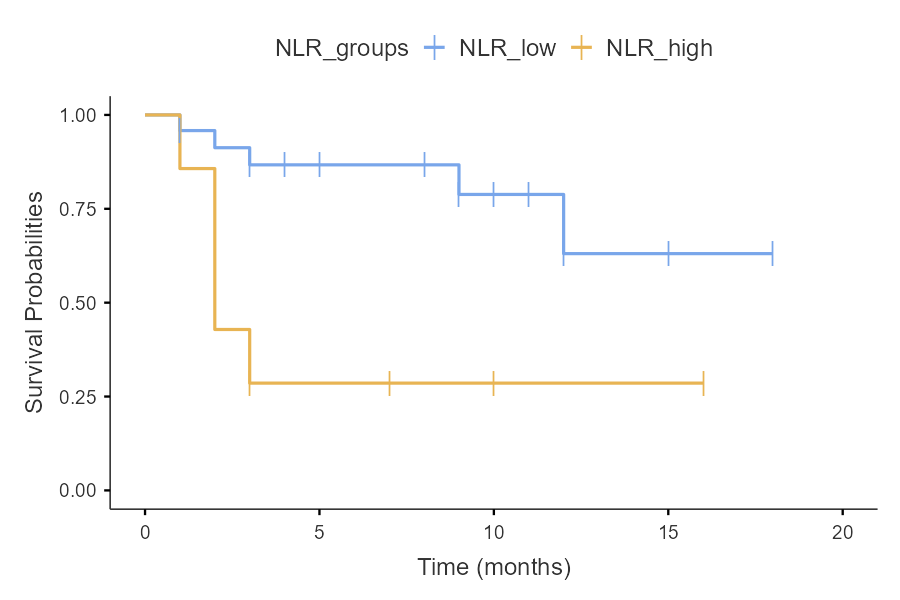
