## Supplementary material for "Neutrophil-to-Lymphocyte Ratio Predicts Infusion-Site Skin Nodules in Parkinson’s Disease Patients Receiving Foslevodopa/Foscarbidopa Subcutaneous Infusion": Tables

### Table 1: Baseline demographic, clinical, infusion-related, and immunological characteristics of the study cohort (N = 38).

| Variable | Mean ± SD, n (%) |
| --- | --- |
| *Baseline demographic and clinical characteristics* | |
| Sex, male | 21 (55.26%) |
| Age (years) | 66.47 ± 7.86 |
| Age at PD onset (years) | 51.45 ± 8.94 |
| Disease duration (years) | 15.03 ± 5.33 |
| BMI (kg/m²) | 22.60 ± 4.62 |
| *Baseline motor and cognitive profile* | |
| UPDRS-III ON | 28.47 ± 13.56 |
| UPDRS-IV | 7.19 ± 2.32 |
| Hoehn & Yahr |  |
| - stage 2 | 16 (42.10%) |
| - stage 2.5 | 9 (23.68%) |
| - stage 3 | 11 (28.95%) |
| - stage 4 | 2 (5.27%) |
| MoCA score | 20.03 ± 5.20 |
| MMSE score | 26.05 ± 3.21 |
| FAB score | 13.65 ± 3.78 |
| *Comorbidities and other relevant therapies* | |
| Diabetes | 1 (2.63%) |
| Use of anticoagulants | 2 (5.26%) |
| Use of antiplatelet agents | 2 (5.26%) |
| *Dopaminergic therapy and infusion-related parameters* | |
| Baseline total LEDD before LDp/CDp-CSI (mg/day) | 1091.13 ± 348.61 |
| DAs use before LDp/CDp-CSI | 22 (57.89%) |
| MAOBIs use before LDp/CDp-CSI | 17 (44.74%) |
| COMTIs use before LDp/CDp-CSI | 17 (44.74%) |
| Amantadine use before LDp/CDp-CSI | 7 (18.42%) |
| Initial foslevodopa LEDD (mg/day) | 953.79 ± 262.46 |
| Duration of infusion, 24 h | 38 (100%) |
| Extra-dose foslevodopa LEDD (mg/day) | 83.21 ± 50.19 |
| Needle length  - 6 mm  - 9 mm | 32 (84.21%)  6 (15.79%) |
| Cannula change interval 24h/48h/72h | 7 (18.42%) /26 (68.42%) /5 (13.16%) |
| Caregiver/patient compliance, good | 32 (84.21%) |
| *Immunological profile* | |
| Neutrophil count (×10⁹/L) | 3.10 ± 0.84 |
| Lymphocyte count (×10⁹/L) | 1.75 ± 0.50 |
| NLR | 1.87 ± 0.59 |

### Abbreviations: *BMI*, Body Mass Index; *UPDRS*, Unified Parkinson’s Disease Rating Scale; *MoCA*, Montreal Cognitive Assessment; *MMSE*, Mini-Mental State Examination; *FAB*, Frontal Assessment Battery; *LEDD*, levodopa equivalent daily dose; *LDp/CDp-CSI*, foslevodopa/foscarbidopa continuous subcutaneous infusion; *DAs*, dopamine agonists; *MAOBIs*, monoamine oxidase B inhibitors; *COMTIs*, Catechol-O-methyltransferase inhibitors; *NLR*, neutrophil-to-lymphocyte ratio.

**Table 2: Comparison between PD-CSN and PD-NSR at 3 months**.

| Variable | PD-CSN (N = 16) | PD-NSR (N = 22) | *P*-value |
| --- | --- | --- | --- |
| *Baseline demographic and clinical characteristics* | | | |
| Sex (male, %) | 9 (56.25%) | 12 (54.54%) | 0.920 |
| Age (years) | 67.06 ± 6.96 | 66.05 ± 8.59 | 0.699 |
| Age at PD onset (years) | 50.81 ± 9.12 | 51.91 ± 8.99 | 0.714 |
| Disease duration (years) | 16.25 ± 5.40 | 14.14 ± 5.23 | 0.233 |
| BMI (kg/m²) | 21.35 ± 4.34 | 23.51 ± 4.70 | 0.158 |
| Time to nodule onset (months) | 1.94 ± 0.77 | - | - |
| *Baseline motor and cognitive profile* | | | |
| UPDRS-III ON | 29.38 ± 14.31 | 27.82 ± 13.28 | 0.732 |
| UPDRS-IV | 7.79 ± 2.15 | 6.82 ± 2.38 | 0.227 |
| Hoehn & Yahr |  |  | 0.137 |
| - stage 2 | 5 (31.25%) | 11 (50%) |  |
| - stage 2.5 | 2 (12.50%) | 4 (18.18%) |  |
| - stage 3 | 7 (43.75%) | 7 (31.82%) |  |
| - stage 4 | 2 (12.50%) | 0 (0%) |  |
| MoCA score | 18.28 ± 5.64 | 21.31 ± 4.57 | **0.040** |
| MMSE score | 25.50 ± 4.02 | 26.46 ± 2.49 | 0.248 |
| FAB score | 12.36 ± 2.97 | 14.60 ± 4.08 | 0.086 |
| *Comorbidities and other relevant therapies* | | | |
| Diabetes | 1 (6.25%) | 0 (0%) | 0.420 |
| Use of anticoagulants | 1 (6.25%) | 1 (4.54%) | 1.0 |
| Use of antiplatelet agents | 1 (6.25%) | 1 (4.54%) | 1.0 |
| *Infusion-related parameters* | | | |
| Baseline total LEDD before LDp/CDp-CSI (mg/day) | 1094.44 ± 392.23 | 1088.73 ± 322.84 | 0.961 |
| Initial foslevodopa LEDD (mg/day) | 1021.70 ± 296.09 | 904.40 ± 229.53 | 0.236 |
| Foslevodopa LEDD at 3-month follow-up (mg/day) | 1105.43 ± 258.50 | 977.04 ± 304.59 | 0.181 |
| Duration of infusion, 24h | 15 (93.75%) | 21 (95.45%) | 1.0 |
| Extra-dose foslevodopa LEDD at 3-month follow-up (mg/day) | 96.69 ± 53.66 | 73.41 ± 46.30 | 0.143 |
| Needle length  - 6 mm  - 9 mm | 15 (93.75%)  1 (6.25%) | 17 (77.27%)  5 (22.73%) | 0.370 |
| Cannula change interval 24h/48h/72h | 4 (25%)/9 (56.25%)/3 (18.75%) | 3 (13.64%)/17 (77.27%)/2 (9.09%) | 0.410 |
| Caregiver/patient compliance, good | 13 (81.25%) | 19 (86.36%) | 0.682 |
| *Immunological profile* | | | |
| Neutrophil count (×10⁹/L) | 3.44 ± 0.90 | 2.85 ± 0.72 | **0.030** |
| Lymphocyte count (×10⁹/L) | 1.67 ± 0.50 | 1.81 ± 0.50 | 0.414 |
| NLR | 2.16 ± 0.61 | 1.65 ± 0.48 | **0.007** |

### Abbreviations: *BMI*, Body Mass Index; *UPDRS*, Unified Parkinson’s Disease Rating Scale; *MoCA*, Montreal Cognitive Assessment; *MMSE*, Mini-Mental State Examination; *FAB*, Frontal Assessment Battery; *LEDD*, levodopa equivalent daily dose; *LDp/CDp-CSI*, foslevodopa/foscarbidopa continuous subcutaneous infusion; *NLR*, neutrophil-to-lymphocyte ratio.

Significant *P*-values are highlighted in bold.

**Table 3: ANCOVA results of inflammatory markers according to nodule status.**

| Time point | Outcome | Model | F (nodule status) | η_p_^2^ | *P*-value | Sensitivity analysis  (*P*-value) |
| --- | --- | --- | --- | --- | --- | --- |
| 3 months | **Neutrophil count** | Principal | 10.856 | 0.259 | **0.002** | **0.003** |
|  |  | Exploratory (with LEDD) | 9.092 | 0.233 | **0.005** | – |
|  | **NLR** | Principal | 7.063 | 0.186 | **0.012** | **0.049** |
|  |  | Exploratory (with LEDD) | 5.366 | 0.152 | **0.028** | – |
| Last follow-up | **Lymphocyte count (Model 1)** | Principal | 6.773 | 0.220 | **0.016** | 0.073 |
|  |  | Exploratory (with LEDD) | 5.538 | 0.187 | **0.027** | – |
|  | **Lymphocyte count (Model 2)** | Principal | 8.753 | 0.267 | **0.007** | **0.019** |
|  |  | Exploratory (with LEDD) | 8.378 | 0.267 | **0.008** | – |
|  | **NLR (Model 1)** | Principal | 18.168 | 0.431 | **<0.001** | **0.002** |
|  |  | Exploratory (with LEDD) | 15.518 | 0.403 | **0.001** | – |
|  | **NLR (Model 2)** | Principal | 12.694 | 0.346 | **0.002** | **0.007** |
|  |  | Exploratory (with LEDD) | 13.837 | 0.376 | **0.001** | – |

Table notes:

Principal models (3-month) adjusted for age, sex, MoCA score, and BMI.

Principal models (last follow-up): Model 1 adjusted for age, sex, BMI; Model 2 adjusted for age, sex, FAB score.

Exploratory models additionally included foslevodopa LEDD at the corresponding time point to test for pharmacological confounding. Sensitivity analyses were performed on principal models and excluded two patients on antiplatelet therapy.

η_p_^2^: partial eta squared. Significant *P*-values are highlighted in bold.

### Table 4: Comparison between PD-CSN and PD-NSR at last follow-up (≥ 6 months).

| Variable | PD-CSN (N = 12) | PD-NSR (N = 18) | *P*-value |
| --- | --- | --- | --- |
| *Baseline demographic and clinical characteristics* | | | |
| Sex (male, %) | 5 (41.67%) | 11 (61.11%) | 0.296 |
| Age (years) | 66.92 ± 7.01 | 64.56 ± 8.00 | 0.411 |
| Age at PD onset (years) | 51.00 ± 10.02 | 50.00 ± 9.33 | 0.782 |
| Disease duration (years) | 15.92 ± 5.71 | 14.56 ± 5.14 | 0.502 |
| BMI (kg/m²) | 20.13 ± 4.61 | 25.03 ± 4.37 | **0.006** |
| Time to nodule onset (months) | 3.50 ± 3.40 | - | **-** |
| Duration of follow-up (months) | 12.17 ± 5.11 | 10.22 ± 3.33 | 0.418 |
| *Motor and cognitive profile (baseline and last follow-up)* | | | |
| UPDRS-III ON (baseline) | 29.50 ± 14.06 | 28.94 ± 15.18 | 0.920 |
| UPDRS-III ON (follow-up) | 21.08 ± 9.69 | 22.47 ± 9.46 | 0.703 |
| Improvement UPDRS-III (%) | 29.58 ± 17.75 | 14.39 ± 53.59 | 1.0 |
| UPDRS-IV (baseline) | 7.73 ± 2.05 | 6.00 ± 2.24 | 0.051 |
| UPDRS-IV (follow-up) | 4.91 ± 1.87 | 4.24 ± 2.05 | 0.387 |
| Improvement UPDRS-IV (%) | 36.37 ± 18.29 | 29.28 ± 29.99 | 0.654 |
| Hoehn & Yahr (baseline) |  |  | 0.104 |
| - stage 2 | 3 (25%) | 9 (50%) |  |
| - stage 2,5 | 2 (16.67%) | 4 (22.22%) |  |
| - stage 3 | 6 (50%) | 5 (27.78%) |  |
| - stage 4 | 1 (8.33%) | 0 (0%) |  |
| MoCA score | 18.68 ± 5.65 | 21.13 ± 4.96 | 0.077 |
| MMSE score | 25.51 ± 4.73 | 26.28 ± 2.71 | 0.882 |
| FAB score | 12.00 ± 3.21 | 14.97 ± 4.06 | **0.042** |
| *Comorbidities and other relevant therapies* | | | |
| Diabetes | 1 (8.33%) | 0 (0%) | 0.400 |
| Use of anticoagulants | 0 (0.00%) | 0 (0%) | - |
| Use of antiplatelet agents | 1 (8.33%) | 1 (5.56%) | 0.765 |
| *Infusion-related parameters (baseline and last follow-up)* | | | |
| Baseline total LEDD before LDp/CDp-CSI (mg/day) | 1049.25 ± 419.59 | 1125.61 ± 287.83 | 0.558 |
| Initial foslevodopa LEDD (mg/day) | 958.80 ± 325.86 | 946.71 ± 225.95 | 0.751 |
| Foslevodopa LEDD at last follow-up (mg/day) | 1064.20 ± 311.54 | 1036.62 ± 315.78 | 0.641 |
| Total LEDD at last follow-up (mg/day) | 1232.78 ± 339.20 | 1362.34 ± 558.69 | 0.703 |
| Duration of infusion, 24h | 11 (91.67%) | 18 (100%) | 0.400 |
| Extra-dose foslevodopa LEDD at last follow-up (mg/day) | 96.33 ± 58.03 | 78.39 ± 46.24 | 0.355 |
| Needle length  - 6 mm  - 9 mm | 11 (91.67%)  1 (8.33%) | 13 (72.22%)  5 (27.78%) | 0.358§ |
| Cannula change interval 24h/48h/72h | 4 (33.33%)/7 (58.33%)/ 1 (8.33%) | 2 (11.11%)/13 (72.22%)/3 (16.67%) | 0.458 |
| Caregiver/patient compliance, good | 10 (83.33%) | 17 (94.44%) | 0.163 |
| *Immunological profile* | | | |
| Neutrophil count (×10⁹/L) | 3.22 ± 0.88 | 3.02 ± 0.84 | 0.529 |
| Lymphocyte count (×10⁹/L) | 1.38 ± 0.31 | 1.97 ± 0.50 | **0.002** |
| NLR | 2.35 ± 0.53 | 1.60 ± 0.51 | **0.001** |

### Abbreviations: *BMI*, Body Mass Index; *UPDRS*, Unified Parkinson’s Disease Rating Scale; *MoCA*, Montreal Cognitive Assessment; *MMSE*, Mini-Mental State Examination; *FAB*, Frontal Assessment Battery; *LEDD*, levodopa equivalent daily dose; *LDp/CDp-CSI*, foslevodopa/foscarbidopa continuous subcutaneous infusion; *NLR*, neutrophil-to-lymphocyte ratio.

Significant *P*-values are highlighted in bold.
